# The Inequality of Neural Destiny: Signatures of Life course Socioeconomic Conditions in Brain Myelination and Grey Matter Volume

**DOI:** 10.1101/2020.06.04.20121913

**Authors:** Leyla Loued-Khenissi, Olga Trofimova, Peter Vollenweider, Pedro Marques-Vidal, Martin Preisig, Antoine Lutti, Matthias Kliegel, Carmen Sandi, Ferhat Kherif, Silvia Stringhini, Bogdan Draganski

## Abstract

Socioeconomic status (SES) plays a significant role in health and disease. At the same time, early-life conditions affect neural function and structure, suggesting the brain may be a conduit for the biological embedding of SES. Here, we investigate the neural signatures of SES in a large-scale population cohort aged 45–85 years. We assess both grey matter volume (GMV) and magnetization transfer (MT) saturation, indicative of myelin content. Higher SES in childhood and adulthood associated with more GMV in several brain regions, including postcentral and temporal gyri, cuneus, and cerebellum, while low SES correlated with larger entorhinal cortex volume. High childhood SES was linked to more widespread GMV differences and higher myelin content in the sensorimotor network while low SES correlated to myelin content in the temporal lobe. Crucially, childhood SES differences in adult brains persisted even after controlling for adult SES, highlighting the unique contribution of early-life conditions to neural status in older age, independent of later changes in SES. These findings inform on the biological underpinnings of social inequality, particularly as it pertains to early-life conditions.

## Introduction

Low socio-economic status (SES) contributes to negative health outcomes ((*1*)), including cardiovascular disease ((*2*)), diabetes ((*3*)) and decreased life expectancy ((*4*)). SES is further linked to differences in cognitive function ((*5*)). For instance, an increased risk of dementia is observed among disadvantaged socioeconomic groups ((*6*)), underscoring the putative link between brain health and SES ((*7*)). Evidence points to a cumulative effect of socio-economic disadvantage over time on health outcomes ((*8*)) highlighting the need to adopt a lifecourse perspective when probing links between SES and physiological markers of health.

Links between SES and cognition suggest the brain is a plausible candidate for the biological embedding of SES. In the developing brain, childhood SES is tied to anatomy ((*9*)) and function ((*10*)), such as reading abilities ((*11*)). Specifically, hippocampal volumes correlate positively with SES ((*12*), (*13*)), as does cortical thickness ((*14*)). These observations suggest childhood SES may mediate effects on language ((*15*)); reading abilities ((*16*)) and mental health status ((*17*)). Studies in adults remain limited, but also highlight links between SES and regional brain volumes, especially in memory regions such as the hippocampus ((*18*), (*19*), (*20*), (*21*)), although a recent meta-analysis highlighted the diversity of specific SES neural correlates across studies ((*22*)).

Studies on the neural imprints of SES nevertheless remain comparatively sparse ((*23*)) and at times yield varied results ((*24*), (*22*)). Further, research to date has relied on region-of-interest (ROI) analyses rather than a whole-brain investigation, leaving results open to bias ((*25*)). This tendency to limit analysis to specific regions may reflect the nature of the feature studied: SES presents a social construct, as opposed to a nosological entity, and therefore corollary neural differences in the population should be subtle, requiring large-scale studies to be identified at the whole-brain level.

It further remains unclear whether SES-mediated differences in late-life reflect traces of childhood SES, as the latter may resolve with a higher SES in adulthood or maturation, or, conversely, persist into old age. To date, few have queried the human brain to assess distal, neural traces of economic conditions in childhood ((*26*)). While some studies uncover an association between childhood SES and increased hippocampal volumes in adulthood ((*27*)), others do not ((*18*), (*28*)). Furthermore, most studies assess grey matter measures to detect exogenously mediated neural differences, but it is white matter that is more susceptible to plastic changes in adulthood ((*29*)) and therefore especially pertinent to neural correlates of social adversity. While some have sought SES-mediated white matter differences in children ((*30*)) and adults ((*31*)), they have primarily employed diffusion-tensor imaging. Tensor-based measures of white matter microstructure lack a straightforward neuro-biological interpretation ((*32*)) and notably are not site-invariant ((*33*)). Non-invasive in vivo white matter assessment remains a challenging endeavor ((*34*)) but magnetization-transfer (MT) saturation quantitative MRI (qMRI) offers a reliable marker of myelin content ((*35*), (*36*)). MT refers to the magnetization exchange between free protons and those bound to macromolecules such as myelin ((*37*)). Imaging studies have found MT quantities correlate to ex-vivo histological assessment of myelin in post-mortem brains ((*38*)), and in addition, have the added benefit of being site-invariant ((*39*)). MT’s enhanced myelin sensitivity may better serve in highlighting white matter variation in a healthy population, where differences are expected to be small.

In this study, we probe the relationship between brain microstructural properties, specifically grey matter volume and myelin content, and SES in a population cohort of older adults, using quantitative MRI ((*40*)). We hypothesize that childhood SES will be reflected in late-life neural markers even when adjusting for SES in adulthood. We investigate these differences at both the whole-brain level, to query differences that may be evoked by qMRI’s sensitivity; and also examine a set of a priori regions, notably the hippocampus and associated memory areas. We query this hypothesis by analyzing a large population cohort (n = 1166) of older adults (mean age = 59.65 years) from one scanner site; employing quantitative neuroimaging using multiparametric maps; applying a data-driven measure of SES; and exploiting a reliable marker of myelin. We hypothesize that differences will be observed in both grey matter and white matter, as quantified by MT saturation mapping.

## Results

Brain imaging data included whole-brain maps of MT saturation and grey matter volume (GMV). MT saturation in grey matter (MTGM) and white matter (MTWM) was analyzed in separate statistical models. Thus, three different sets of brain anatomy features constituted our outcome measures in a general linear model (GLM). We designed three multiple regression tests in SPM to examine brain differences linked to SES in the cohort for each neuroimaging data set. In the first two, we included either adult SES (aSES) or childhood SES (cSES) as a covariate of interest. For the third, we designed a full model that included both aSES and cSES. By combining the two SES variables, we can assess the unique contribution of one or the other to neural outcome variables. Importantly, we did not orthogonalize these two measures as no firm principle can attribute primacy to one or the other. Finally, age, gender and total intracranial volume (TIV) – a proxy for head size – were included in the design as nuisance variables ((*41*)). We tested for the overall contribution of SES to differences in neural data by performing a one-sample t-test on the SES coefficient of the model. Mass-univariate analyses were performed with a significance threshold of p = 0.05, FWE corrected for multiple comparisons at the whole-brain voxel level ((*42*)).

## Results for SES

### Model 1 – Brain changes associated with Adult SES

#### MT changes associated with Adult SES

Adult SES was tied to decreases in MT density bilaterally in the entorhinal cortex. (Figure 2)

**Figure 1:**
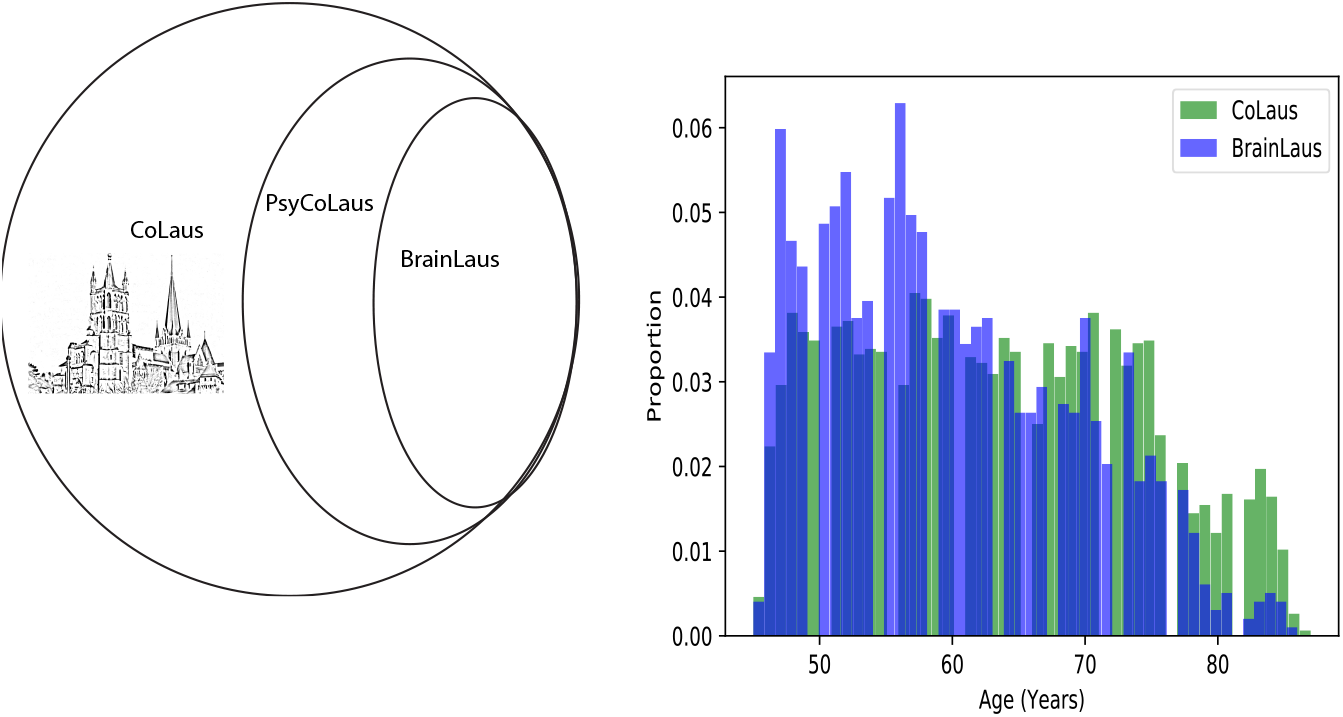
Cohort Characteristics. A) The BrainLaus study comprises a subset of the PsyCoLaus cohort, which is itself a subset of the population cohort (Cohorte Lausanne, CoLaus). B) The CoLaus Cohort includes a representative sample of the population, which is reflected in the BrainLaus subset, but for age. Here, age distributions for participants in the BrainLaus study are shown alongside age distributions for participants that did not undergo MR scanning.

**Figure 2:**
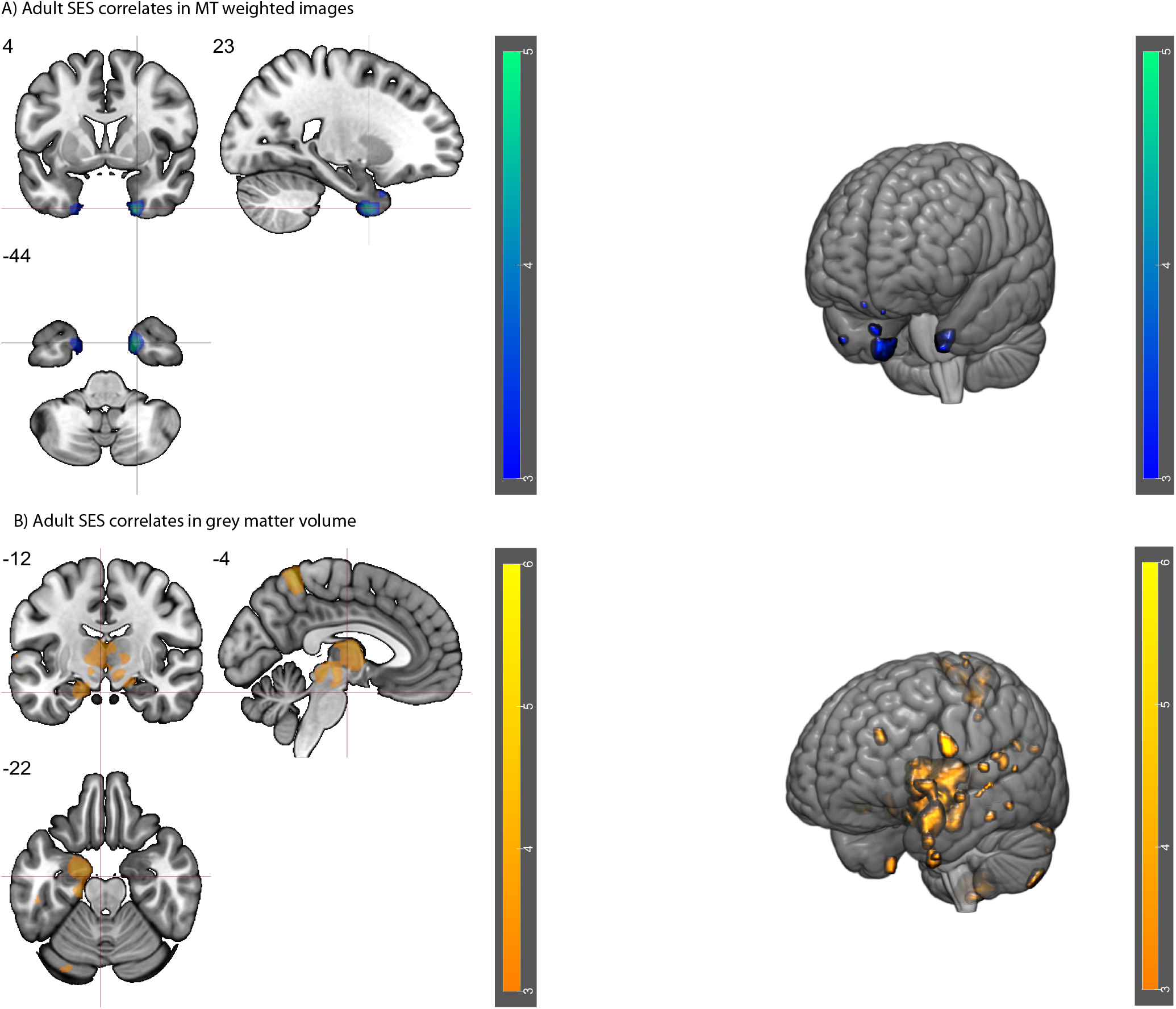
Results of GLM 1 (adult SES). A) Results of a negatively signed one-sample t-test of adult SES on MT in grey matter. B) Results of a positively signed one-sample t-test of adult SES on grey matter volume. Colorbars indicate t-values. All maps shown are thresholded at p = 0.05, FWE corrected.

#### Grey Matter volume changes associated with Adult SES

Adult SES correlated positively with grey matter volume in several regions, including bilateral superior parietal lobules, left thalamus and right cerebellum (exterior).

### Model 2 – Brain changes associated Childhood SES

#### MT changes associated Childhood SES

Childhood SES (cSES) correlated positively with MT in right superior parietal lobule and negatively with MT in bilateral temporal lobes. In white matter, cSES correlated positively with MT near the pallidum/ventral tegmentum; bilateral precentral gyrus; and right inferior occipital gyrus (Table 2). The pattern found in MT notably delineates the sensorimotor network ((*43*)) (Figure 3).

**Figure 3:**
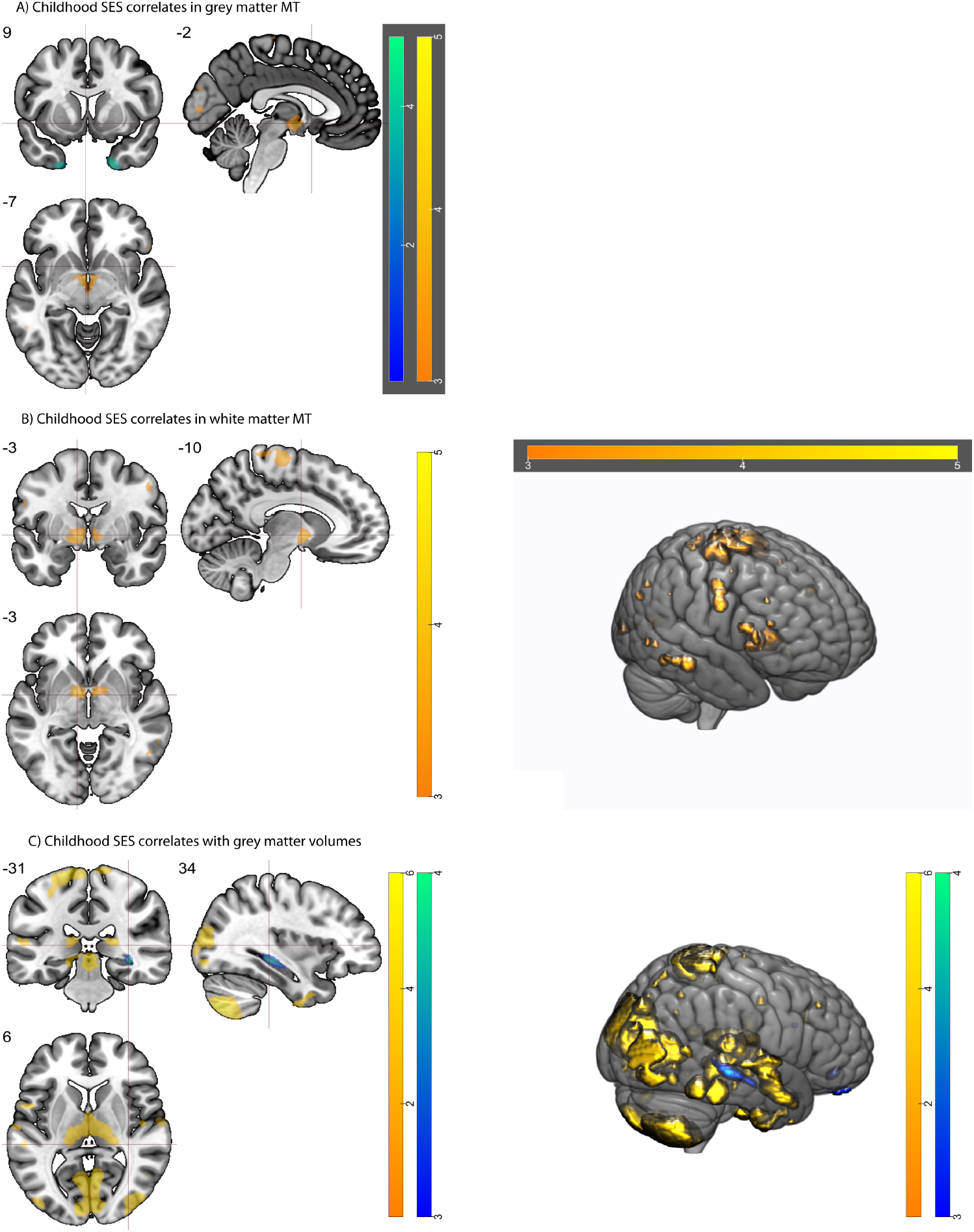
Results of GLM 2 (Childhood SES) in MT maps and grey matter volumes. A) Results of both positive and negatively signed one-sample t-tests of childhood SES on MT in grey matter. B) Results of a positively signed one-sample t-test on childhood SES on MT in white matter. C) Results of both positive and negatively signed one-sample t-tests on childhood SES on in grey matter. Colorbars indicate t-values (hot for positive and cold for negative t-tests). All maps shown are thresholded at p = 0.05, FWE corrected.

**Table 1:**
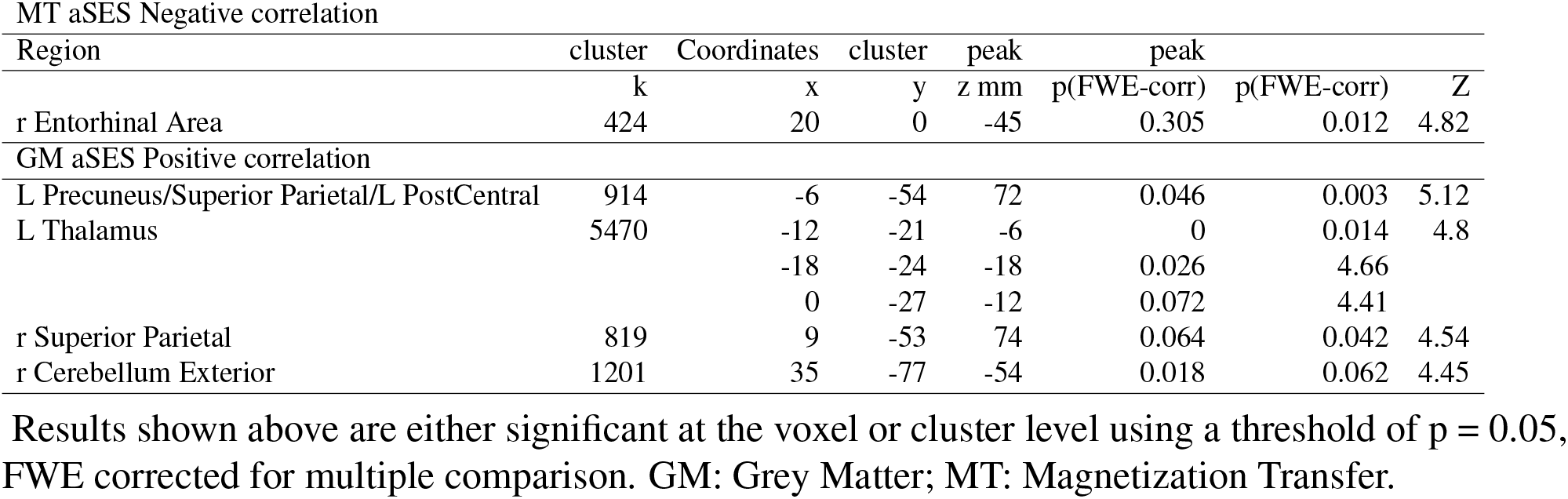
Whole brain voxel level analysis of MT load and grey matter volumes in relation to adult SES (aSES).

**Table 2:**
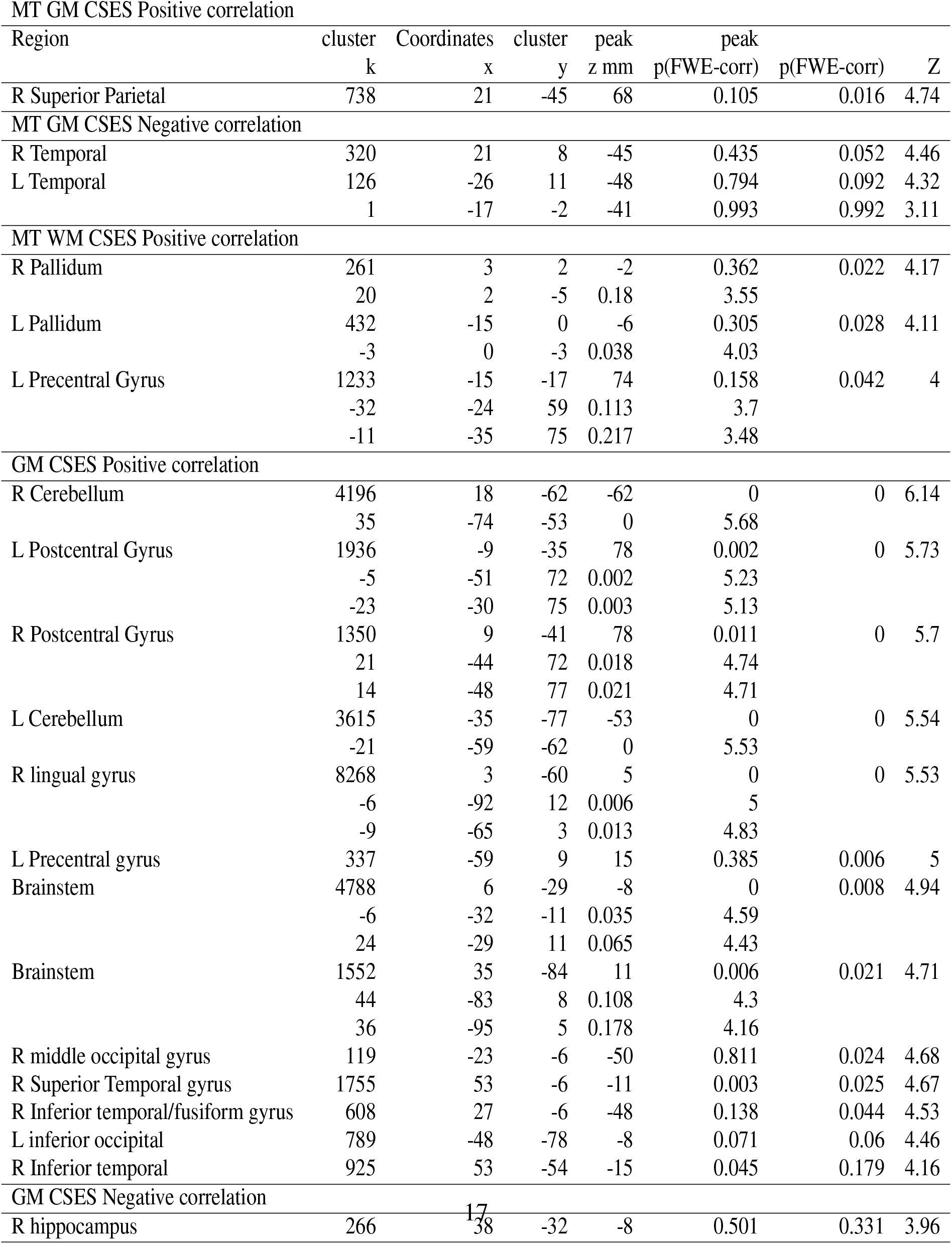
Whole brain voxel level analysis of MT load and grey matter volumes in relation to childhood SES (cSES).

**Table 3:**
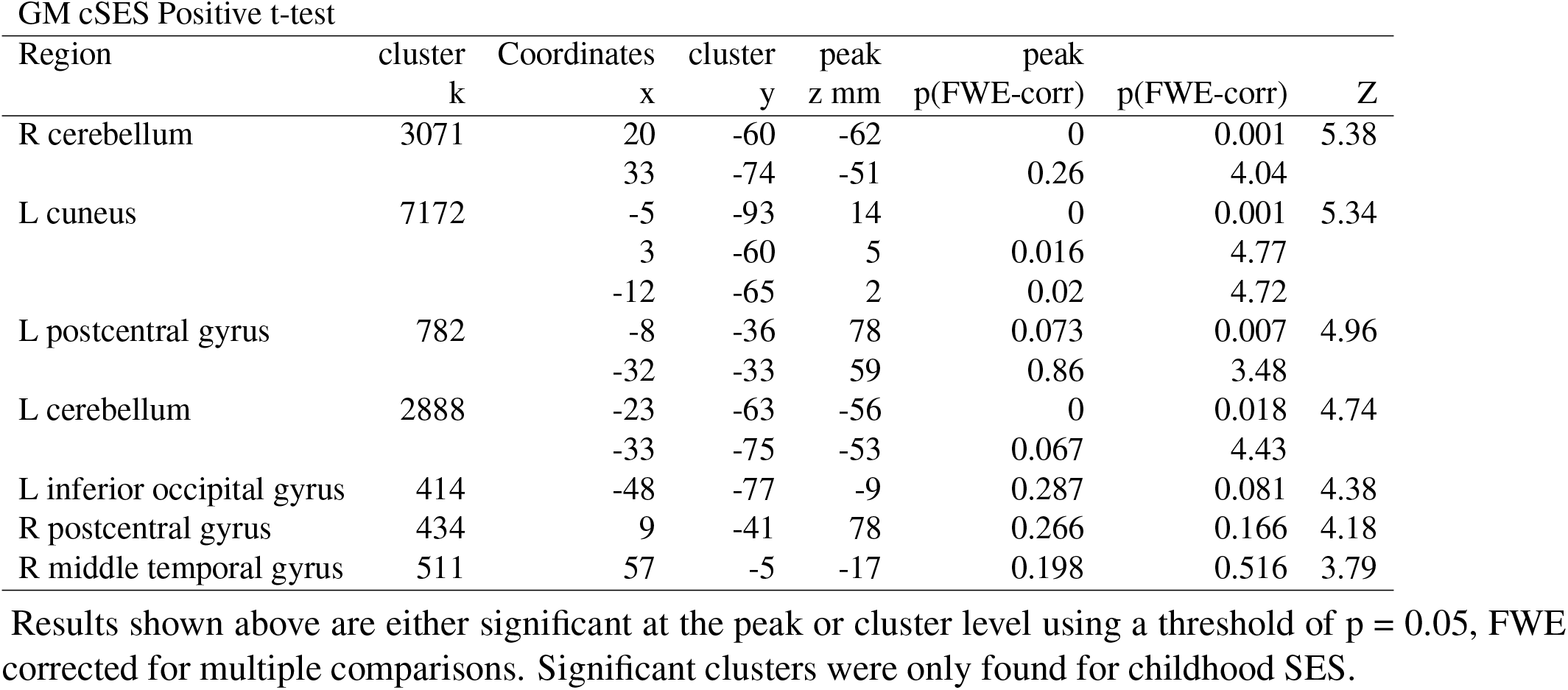
Whole brain voxel level analysis of grey matter volumes in relation to SES in a full model including both childhood and adult SES.

| GM cSES Positive t-test |  |  |  |  |  |  |  |
| --- | --- | --- | --- | --- | --- | --- | --- |
| Region | cluster<br>k | Coordinates<br>x | cluster<br>y | peak<br>z mm | peak<br>p(FWE-corr) | peak<br>p(FWE-corr) | Z |
| R cerebellum | 3071 | 20 | -60 | -62 | 0 | 0.001 | 5.38 |
|  |  | 33 | -74 | -51 | 0.26 | 4.04 |  |
| L cuneus | 7172 | -5 | -93 | 14 | 0 | 0.001 | 5.34 |
|  |  | 3 | -60 | 5 | 0.016 | 4.77 |  |
|  |  | -12 | -65 | 2 | 0.02 | 4.72 |  |
| L postcentral gyrus | 782 | -8 | -36 | 78 | 0.073 | 0.007 | 4.96 |
|  |  | -32 | -33 | 59 | 0.86 | 3.48 |  |
| L cerebellum | 2888 | -23 | -63 | -56 | 0 | 0.018 | 4.74 |
|  |  | -33 | -75 | -53 | 0.067 | 4.43 |  |
| L inferior occipital gyrus | 414 | -48 | -77 | -9 | 0.287 | 0.081 | 4.38 |
| R postcentral gyrus | 434 | 9 | -41 | 78 | 0.266 | 0.166 | 4.18 |
| R middle temporal gyrus | 511 | 57 | -5 | -17 | 0.198 | 0.516 | 3.79 |

Results shown above are either significant at the peak or cluster level using a threshold of $p = 0.05$ , FWE corrected for multiple comparisons. Significant clusters were only found for childhood SES.

**Table 4:**
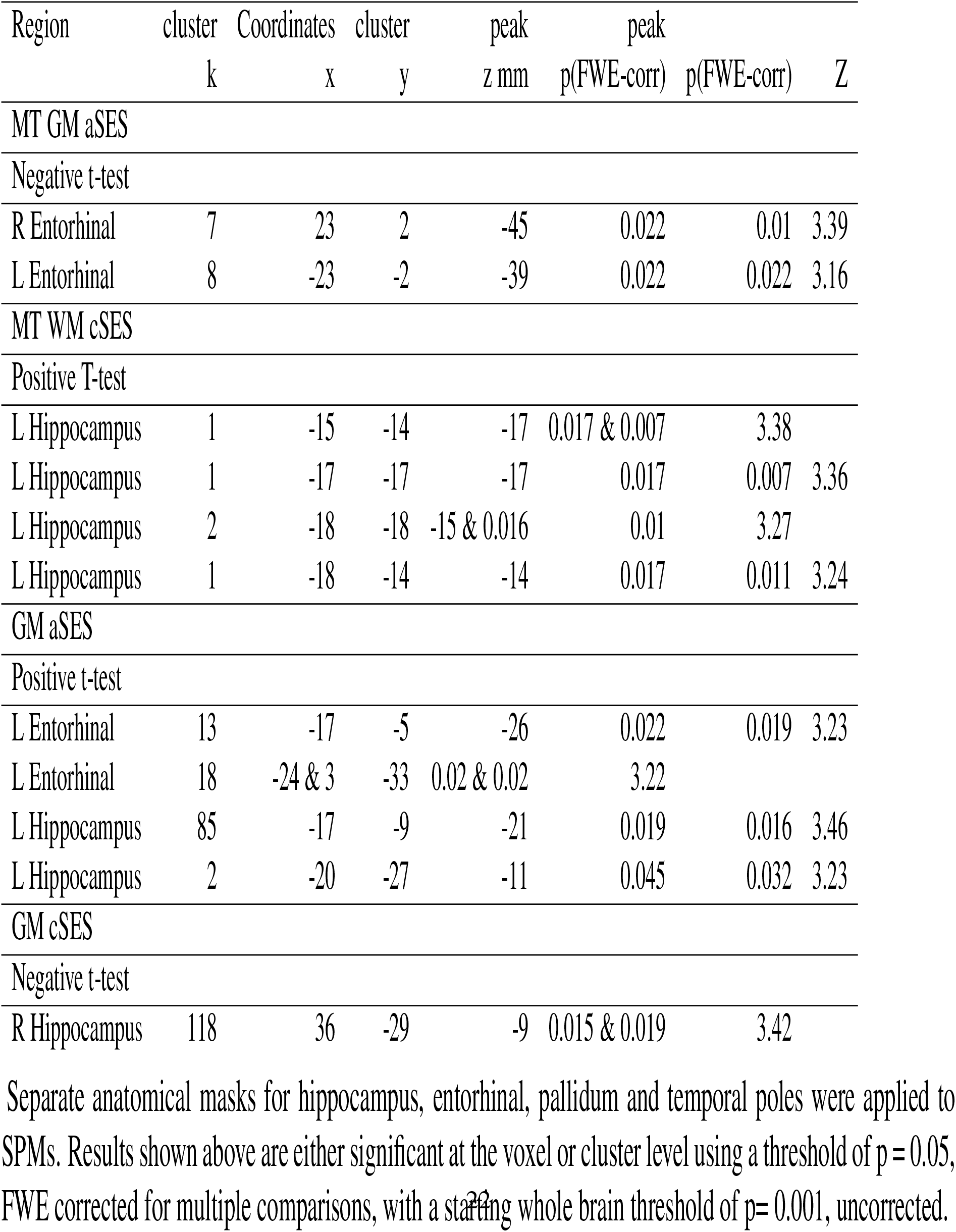
Small volume correction analysis of MT load and grey matter volumes in relation to childhood and adult SES in the full model.

#### Grey Matter volume changes associated Childhood SES

Childhood SES correlated positively with grey matter volume in several regions, including bilateral cerebellum and postcentral gyri, right lingual gyrus, brainstem, left pre-central gyrus, right middle occipital gyrus, right superior temporal gyrus, left inferior occipital gyrus and the inferior temporal gyri bilaterally. Using small volume correction, we also find a negative correlation between childhood SES and right hippocampus volume.

#### GLM 3 – Full model

Results above suggest childhood SES has a stronger relationship with late-life adult brain anatomy than does adult SES. To better inform our hypothesis, we further analyzed both child- and adulthood SES in the same model. Because these two variables are correlated (r = 0.536, p ¡ 0.001), we do not expect associated regression slopes to survive thresholds corrected for multiple tests at the whole brain level. Below, we report either peaks or clusters whose t-values fall below a threshold of p = 0.05, FWE corrected. Based on the extant literature, we use a small volume correction for a set of memory regions including: the parahippocampal gyrus, entorhinal cortex and the hippocampus. At the whole brain level, significant results were found exclusively for positive associations between childhood SES and grey matter in bilateral cerebellum; left cuneus; and left postcentral gyrus (Figure 4).

**Figure 4:**
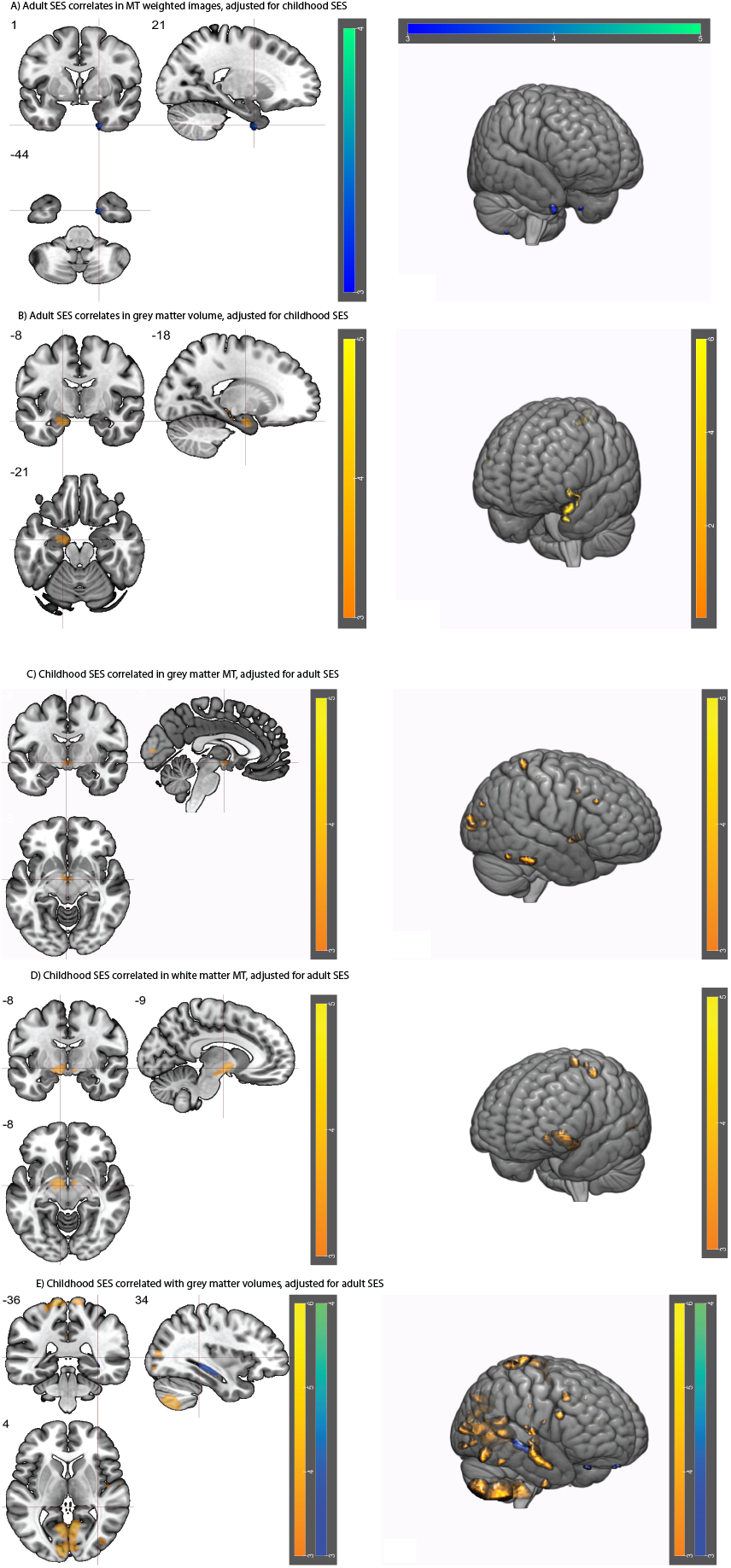
Results of GLM 3 including adult and childhood SES as covariates in MT maps and grey matter volumes. A) Results of a negatively signed one-sample t-test of adult SES in MT in temporal pole; B) Results of a positively signed one-sample t-test on adult SES in entorhinal grey matter volumes; C) and D) results of positive childhood SES correlates in globus pallidus MT (grey and white matter) E) Results of both positive and negative signed one-sample t-tests on childhood SES in grey matter. Colorbars indicate t-values. All maps shown are thresholded at p = 0.001, uncorrected for visualization purposes.

#### Small Volume Correction Analysis – Adult SES

Adult SES correlated negatively with MT bilaterally in the entorhinal cortex. Grey matter volumes correlated positively with the left entorhinal cortex and hippocampus.

#### Small Volume Correction Analysis – Childhood SES

Childhood SES correlated positively with MT in white matter in left hippocampus. Childhood SES further correlated negatively with the right hippocampus.

#### GLM 3 – Full model, controlling for cardio-vascular risk factors

Studies on SES commonly take into consideration other health factors when assessing outcomes, though the latter tend towards easily quantifiable variables such as morbidity, as opposed to in-vivo brain measures. We nonetheless opted to perform a GLM that included a regressor for cumulative cardiovascular risk factors, including previous cardiac events; diagnosis of diabetes; and obesity. At the whole brain level, significant results were found only for MT values in a small cluster (k = 13) of the right cuneus when associated with childhood SES.

## Discussion

In this study, we examined the relationship between life-course socioeconomic status (SES) and structural brain properties using MRI-derived estimates indicative of myelin content and grey matter volume in older individuals from the general population in a whole-brain analysis. In contrast to previous studies, we adopted a life-course perspective and hypothesized that neural traces of childhood SES remain when controlling for adult SES. We found that both childhood and adult SES separately correlated with grey matter volume and myelin differences. Moreover, the effect of childhood SES on grey matter volume and myelin content was independent of adult socioeconomic circumstances. Though we find evidence of an effect of adult SES in both grey matter and MT when controlling for childhood SES, childhood SES was associated with more robust neural differences when controlling for adult SES. Our results support the hypothesis that childhood SES leaves a neural imprint even in adulthood and more generally, corroborate the latent effect model for the impact of childhood SES on adult outcomes ((*44*)).

Our findings were uncovered in the largest study to date investigating SES associations with the brain. Studies on neural imprints of SES have yielded variable results, as highlighted in a recent meta-analysis ((*22*)), which may be due in part to limited sample sizes. Two key studies have attempted to overcome this problem ((*13*), (*23*)), finding, in the first, a positive correlation between parental education and hippocampal volume, without adjusting for family income, and an association between cortical surface area and both parental income and education. In the second study, widespread cortical surface area and hippocampus similarly correlate with higher SES, as assessed by the Hollingshead score. While these two studies yield concordant results, they are both found in pediatric cohorts and further target the hippocampus for volumetric analysis. Our whole brain approach finds hippocampal GM associates with lower childhood SES, but higher adult SES in an ROI analysis, appearing to contradict the relationship between the hippocampus and SES. However, qMRI may highlight specificity in this same relationship, as hippocampal volume may cede to increases in myelination ((*36*)). Further, myelin may be a more pertinent metric for function across the lifespan ((*45*), (*46*)).

Scanner site variability((*47*)) may also contribute to differences in neuroimaging studies. An innovative feature of our study is the use of MT saturation maps to extract estimates of grey matter volumes ((*48*)) and myelin content (*35*)). Crucially, quantitative MRI maps are impervious to inter-site variability ((*49*)). Our results therefore offer an added reliability over findings obtained with traditional MRI methods, particularly with regards to myelin quantification methods.

### SES differences in brain’s myelin

Most studies on in vivo structural brain properties linked to SES focus on grey matter volume or cortical thickness measures. However, myelin plays a crucial role in brain function and dysfunction ((*29*)) and therefore should not be neglected. Here, we index myelin with the use of qMRI ((*50*), (*51*)). As shown in our study, MT values covaried with SES in regions distinct from grey matter volume changes, highlighting myelin’s independent status in the brain. Our results support a recent study showing a relationship between neighborhood deprivation, and rate of myelination, as assessed by MT, across childhood and adolescence ((*45*)). Further, we find a positive association of myelin in regions comprising the sensorimotor network with SES. This network has previously been associated to cognitive impairment ((*52*)) and MT in old age correlated with motor performance ((*53*)). Aging induces cognitive decline ((*54*)), even in early adulthood ((*55*)) as well as decreases in motor performance ((*56*)). Converging evidence highlights the increasing association between cognitive and sensorimotor functions with aging ((*57*)). One possibility is that childhood SES may provide a buffer to functional decline in old age via increased myelination of the sensorimotor network.

### Regional Specificity of SES Neural Differences

Brain regions that we found to covary with SES play important function roles in cognition, memory and motor function. The pallidum plays a role in reward and motivation ((*58*), as well as motor function ((*59*)). The hippocampus plays a significant role in memory ((*60*)). The entorhinal cortex is notably involved in spatial ((*61*)) and temporal ((*62*)) memory and acts as a gateway between the hippocampus and cortical areas ((*63*)). The hippocampus in particular has previously been found implicated in psychosocial adversity ((*64*)) and yet is also known as a region susceptible to plasticity ((*65*)). Finally, the temporal pole has been hypothesized to serve in integrating different streams of perceptual information and also mediates social interaction ((*66*)) and higher order knowledge representation ((*67*)). Differences in any of these structures may therefore have considerable functional implications.

### SES differences and the hippocampus

Differences in hippocampal grey matter volume associated with SES have previously been reported in a number of studies ((*21*), (*27*)). Our results support partially support this relationship, but we detect an inverse relationship between right hippocampal volume and childhood SES. We also find a positive, linear relationship between left hippocampal volume and adult SES, specifically. Childhood SES in our cohort was nonetheless also associated with higher right temporal pole volumes, the latter which forms part of the temporal lobe system, and is uniquely sensitive to age-related decline ((*68*)). Childhood SES further correlates positively with greater MT in left hippocampus, raising the possibility of interplay between myelin and grey matter. For instance, cortical grey matter reduction occurs in healthy adolescence along with an increase of myelination ((*69*)). In the same model, adult SES correlated with increased left entorhinal, parahippocampal and hippocampal volumes; and decreased MT in bilateral entorhinal cortex, here directly showing opposing effects of SES on grey matter and myelin in left entorhinal cortex. Our results thus suggest a more complex interaction in temporal lobe regions in relation to SES, with differential effects of myelin, grey matter and early or late-life SES implicated in disparate neural profiles.

### Limitations of the Study

In our study, childhood SES was assessed using adult recall that is susceptible to faulty memories ((*70*)). Household income in childhood and adulthood are further indexed by different measures and it can be argued that the one for childhood skews towards assessing disadvantage – although this bias may be redressed as retrospective assessments tend to favor a more optimistic view of how things were ((*71*)). We also define SES with a composite measure, which does not identify unique risk factors ((*72*)). Finally, our study design precludes the possibility to control for context beyond SES in early life that can impact neural structure. In spite of these limitations, our results are based on a precious dataset, as not all large-scale neuroimaging data have childhood, or conversely, adult data.

### Conclusions

Our study informs the sparse extant literature on brain correlates of SES. Known associations between childhood adversity and late-life outcomes strongly suggest a causal process embedded in the arrow of time. By highlighting a neurophysiological embedding of childhood SES in old age, our results add credence to this association.

## Methods

### Cohort

The study cohort (BrainLaus) was recruited from the CoLaus—PsyCoLaus general population cohort of the city of Lausanne, Switzerland ((*73*); (*74*)). The BrainLaus study aims to scan participants at two time points, spaced 5 years apart. These two time points represent the 3rd and 4th study time points of the greater CoLaus study. Analyses were performed on imaging data acquired between 2014 and 2018 and represented the first BrainLaus time point. A total of 1332 participants were scanned at the Siemens Prisma 3T scanner of the Department for clinical neuroscience, Lausanne University Hospital. (Figure 1a).

### Cohort Description

The CoLaus—PsyCoLaus study was designed to recruit a representative sample of the population ((*73*)). We sought to determine if the BrainLaus subset differed from the larger cohort on a number of key dimensions by examining differences between Co-Laus—PsyCoLaus participants that were not included in the BrainLaus cohort (n = 5401); and the BrainLaus cohort on all measures available for somatic variables (n = 1309). (A full list of variables can be found in Appendix (A)). There was no significant difference in gender, education level, or last known occupation distributions between the two cohorts. A significant difference in age was found between the two cohorts however, with an average age of 63 for CoLaus—PsyCoLaus participants and 59 for BrainLaus participants (Cohen’s d = 0.4). This result underlines the necessity of including age as a nuisance variable in subsequent neuroimaging analyses. (Figure 1b).

### Neuroimaging Data and Analysis

The scanning protocol included a multi-parameter mapping (MPM) relaxometry protocol ((*75*)) and diffusion-weighted acquisition that was not used in the current study. Approximate total scan duration lasted 45 minutes. Analyses were performed in SPM12 (http://www.fil.ion.ucl.ac.uk/spm/) using Matlab, 2017. Socio-economic data were missing for 18 of the n = 1194 participants whose neuroimaging data were retained. A total of n = 1166 participants (mean age: 59.65 years; 622 females, 544 males) were included in our analyses.

Quantitative MT maps were computed from raw MR images acquired using a multi-echo 3D FLASH (fast low angle shot) at a 1 mm, isotropic resolution ((*51*)). The MRI data was acquired with T1-, PD- and MT-weighted contrast (respective Repetition Time/Flip Angle (FA) of 23.7ms/21°C, 23.7ms/6°C and 23.7ms/6°C(MT). For the MT-weighted contrast, an off-resonance Gaussian MT saturation RF pulse (4ms, FA = 220°C, 2KHz frequency offset) was applied before non-selective excitation. Multiple echo images were acquired with echo times ranging from 2.2 ms to 19.7 ms (except for the MT-weighted scans where the maximum echo time was 17.2ms, due to the application of the MT saturation pulse). We employed GRAPPA parallel imaging (acceleration factor of 2) in anterior-posterior phase encoding direction and 6/8 partial Fourier acquisitions in the partition direction (left-right). The Multi-parameter Mapping protocol also included the acquisition of MRI data for the mapping of the radio-frequency excitation field B1, instrumental to obtain accurate MRI biomarkers of tissue myelin density ((*39*)). This data was acquired using the technique described in ((*76*)). Acquisition settings were identical to those described in ((*75*)).

### Image Pre-processing

Acquired data underwent automated pre-processing in the multi-channel “unified segmentation” Bayesian framework of SPM12 yielding GM and WM probability maps derived from MT and PD* maps. A study specific DARTEL template was created from all individual GM and WM tissue classes ((*77*)) to then apply the derived spatial registration parameters onto grey matter volumes and MT saturation maps and warp these to standard MNI space. Here, we followed the default settings for implementation of an established “weighted-averaging” procedure ((*50*)).

### Image quality assessment

Given the size and average age of the cohort, as well as the plurality of MRI data acquired, a multi-step image quality procedure was applied to our cohort. In a first instance, we computed regional averages for MT, R2* and grey matter volumes for each participant by applying individual inverse deformation fields to anatomical volumes provided by the Neuromorphometrics Atlas (Neuromorphometrics, Inc.), yielding 129 values for each participant, for each dataset. Individual average values falling outside a range of +/− 4 standard deviations from the group mean of a specific brain region were flagged and subsequently excluded from analysis (n = 55). In a second instance, we examined differences between individual GM, WM, and CSF segmentations and corresponding canonical tissue probability maps. We first binarized individual tissue segmentations before conducting this procedure. Resulting images were then vectorized, assigned a value of 1 for all voxels *>* 0 and summed. Participants for whom this total exceeded the group average by 3 standard deviations were subsequently excluded from analysis (n = 31). Finally, we performed a visual inspection of datasets that showed high standard deviations of the R2* parameter in white matter. This index has been shown to exhibit a high correlation with motion history during data acquisition ((*78*)). The criterion for a high standard deviation was set to a conservative cut-off, which flagged approximately 500 potentially problematic datasets. As our cohort tended towards an older population, we expect more movement than average. Therefore, we visually examined these 500 datasets to identify gross movement, physiological anomalies or other artefacts. This visual rating excluded another n = 25 subjects from subsequent analysis. Finally, 4 more participants were found to have been scanned with a different coil and were also excluded from the data analysis pool. These image quality procedures excluded a total of 125 subjects from analysis. (Appendix C)

### SES variables

The CoLaus—PsyCoLaus study collected a wide range of socio-demographic variables. Socio-economic status can be indexed in several ways; however, consensus holds that three observable variables can serve as valid measures of the underlying construct, namely, education, income and occupation levels ((*79*)). CoLaus—PsyCoLaus demographic data included information on mean income (in 6 intervals); education (3 levels); and self and partner’s last known occupation. The latter were ranked according to the European Socio-Economic Classification (ESEC) scale (https://www.iser.essex.ac.uk/archives/esec/ user-guide) (9 levels) and assigned a corresponding numerical value; own income was taken to be highest household income between spouses, where applicable. Measures of childhood SES included father’s occupation (ranked according to the ESEC scale); highest parental education (3 levels); and a measure of childhood household financial status as proxy of childhood income (Appendix B). This last measure included a sum of 9 positive and negative answers for family lifestyle and conditions, such as ownership of a car and having insufficient heating. The following variables were scaled into tertiles and assigned values ranging from 0 to 2: Adult occupation, taken as highest household occupation; mean income; paternal occupation; and childhood finances. To obtain a precise measure of adult and childhood SES constructs, we performed two PCAs for the trio of adult and childhood SES variables. We found that, in adulthood, education explained most of the variance (63.7%), followed by income (21.84%) and occupation (14.5%). In childhood, household income explained most of the variance (70.39%), followed by education (16%) and occupation (13.6%). We then created a composite measure of adult SES and one of childhood SES by weighting tertile measures of income, education and occupation with their respective variance contributions before summing them. This procedure allowed for the range of possible SES variables to increase from 3 to 48, with a concomitant increase in information, as formalized by entropy, from 1.58 to 4.68 and 1.55 to 3.67 bits, for adult and childhood SES, respectively. This procedure therefore produces a single, precise measure of SES to include as an independent variable in our analyses.

## Data Availability

De-identified MRI features that were used for the study can be provided upon request. Non-identifiable individual-level data are available for researchers who seek to answer questions related to health and disease in the context of
research projects who meet the criteria for data sharing by research committees. Please follow the instructions at https://www.colaus-psycolaus.ch/ for information on how to submit an application for gaining access to CoLaus data.

## Acknowledgments

The CoLaus—PsyCoLaus study was and is supported by research grants from GlaxoSmithKline, the Faculty of Biology and Medicine of Lausanne, and the Swiss National Science Foundation (grants Nr. 3200B0 105993, 3200B0 118308, 33CSCO 122661, 33CS30 139468 and 33CS30 14840).LLK and SS were supported by the European Commission(H2020 Lifepath grant No. 633666), and by the Leenaards and Jeantet Foundations. BD is supported by the Swiss National Science Foundation (NCCR Synapsy, project grant Nr. 32003B 135679, 32003B 159780) and the Leenaards Foundation. AL is supported by the Swiss National Science Foundation (project grant Nr. 320030 184784) the Roger De Spoelberch foundation. LREN is very grateful to the Roger De Spoelberch and Partridge Foundations for their generous financial support.

